# Early and massive testing saves lives: COVID-19 related infections and deaths in the United States during March of 2020

**DOI:** 10.1101/2020.05.14.20102483

**Authors:** James B. Hittner, Folorunso O. Fasina, Almira L. Hoogesteijn, Renata Piccinini, Prakasha Kempaiah, Stephen D. Smith, Ariel L. Rivas

## Abstract

To optimize epidemiologic interventions, predictors of mortality should be identified. The US COVID-19 epidemic data −reported up to 3-31-2020− were analyzed using kernel regularized least squares regression. Six potential predictors of mortality were investigated: (i) the number of diagnostic tests performed in testing week I; (ii) the proportion of all tests conducted during week I of testing; (iii) the cumulative number of (test-positive) cases through 3-31-2020, (iv) the number of tests performed/million citizens; (v) the cumulative number of citizens tested; and (vi) the apparent prevalence rate, defined as the number of cases/million citizens. Two metrics estimated mortality: the number of deaths and the number of deaths/million citizens. While both expressions of mortality were predicted by the case count and the apparent prevalence rate, the number of deaths/million citizens was ≈3.5 times better predicted by the apparent prevalence rate than the number of cases. In eighteen states, early testing/million citizens/population density was inversely associated with the cumulative mortality reported by 31 March, 2020. Findings support the hypothesis that early and massive testing saves lives. Other factors –e.g., population density–may also influence outcomes. To optimize national and local policies, the creation and dissemination of high-resolution geo-referenced, epidemic data is recommended.

## Bullet points

- A multidimensional –numerical, geographic, demographic and temporal–approach that emphasizes interactions is used to identify predictors of COVID-19 related mortality.
- A combinatorial template helps detect the impact of early testing on mortality.
- This rapidly conducted, policy-oriented analysis applies to many geographic scales.

To control a pandemic associated with a substantial mortality –such as COVID-19–, WHO recommends massive testing [1]. In spite of its relevance, the power of testing-related variables to predict mortality has not yet been empirically investigated in this disease.

To predict and identify when and where mortality is likely to occur, at least three types of metrics may be considered, which focus on: (i) cases (counts), (ii) disease prevalence in a specific geographic location and/or time, and (iii) the demographic density of infected locations [2]. However, assessing the actual prevalence of a disease characterized by a substantial number of asymptomatic infections –such as COVID-19– is not possible, unless 100% of the population is tested with a highly sensitive test, repeatedly [3, 4]. Consequently, we use the term *apparent prevalence* to describe the ratio of test-positive cases to all tested individuals. If expressed per million residents, the apparent prevalence can compare different geographical units, e.g., each and all states of the US.

Unfortunately, to conduct comprehensive studies that investigate numerous states, a protracted research program is required. To rapidly provide policy-makers with usable information, here a quasi-real time assessment was designed, which captures both nationwide and state-specific dimensions. Analyzing the epidemic data reported in all 50 states of the USA, during March of 2020 (the month when testing started), we investigated whether testing-related variables –including massive and early testing– predict mortality.

Six variables were assessed as possible predictors of fatalities: (i) the number of diagnostic tests performed in week I of testing; (ii) the proportion of all tests conducted during the first week of testing; (iii) the cumulative number of (test-positive) cases through 3-31-2020, (iv) the number of tests performed/million citizens; (v) the cumulative number of citizens tested; and (vi) the apparent prevalence rate, defined as the number of cases/million citizens. To examine the predictive ability of these variables, we modeled the data using a nonparametric machine learning approach known as kernel regularized least squares (KRLS) regression [5]. To implement the procedure we used the KRLS *R* software package [6]. KRLS is appropriate when linear regression assumptions –such as linearity and additivity– are not met and the precise functional association between the predictors and criterion is unknown.

Because there is no prior knowledge on the use of these composite variables, no pre-established method or criterion was chosen to analyze the data. Instead, recognition of patterns observed after the data were collected was adopted. When distinct patterns were observed –such as L-shaped data distributions [9]–, thresholds were selected to match the upper limit of a data segment linearly distributed so that the intersection of two orthogonal lines would identify three groups of data.

A public source was used to collect the overall US and state-specific data on the COVID-19 pandemic, which was complemented with state-specific population data [7, 8]. All analyses included data from each state of the US (Supplemental Table 1).

The six predictors accounted for 93.5% of the variance in number of deaths and 86.7% of the variance in deaths/million cases (Supplemental Tables 2A, 2B). Of the six predictors, two were statistically significant: cumulative number of confirmed cases and apparent prevalence rate. These two variables were comparable predictors of mortality count. However, for predicting deaths per million citizens, the apparent prevalence rate was a 3.5 times stronger predictor than was the number of confirmed cases (Supplemental Table 2B).

In addition, the number of tests administered during week one of testing/million citizens/population density distinguished three groups of states when the number of deaths/million citizens was the outcome variable (Fig. 1A). Two of these groups exhibited statistically significantly different medians (*p*<0.001, Mann-Whitney test, Fig. 1B).

**Fig. 1.**
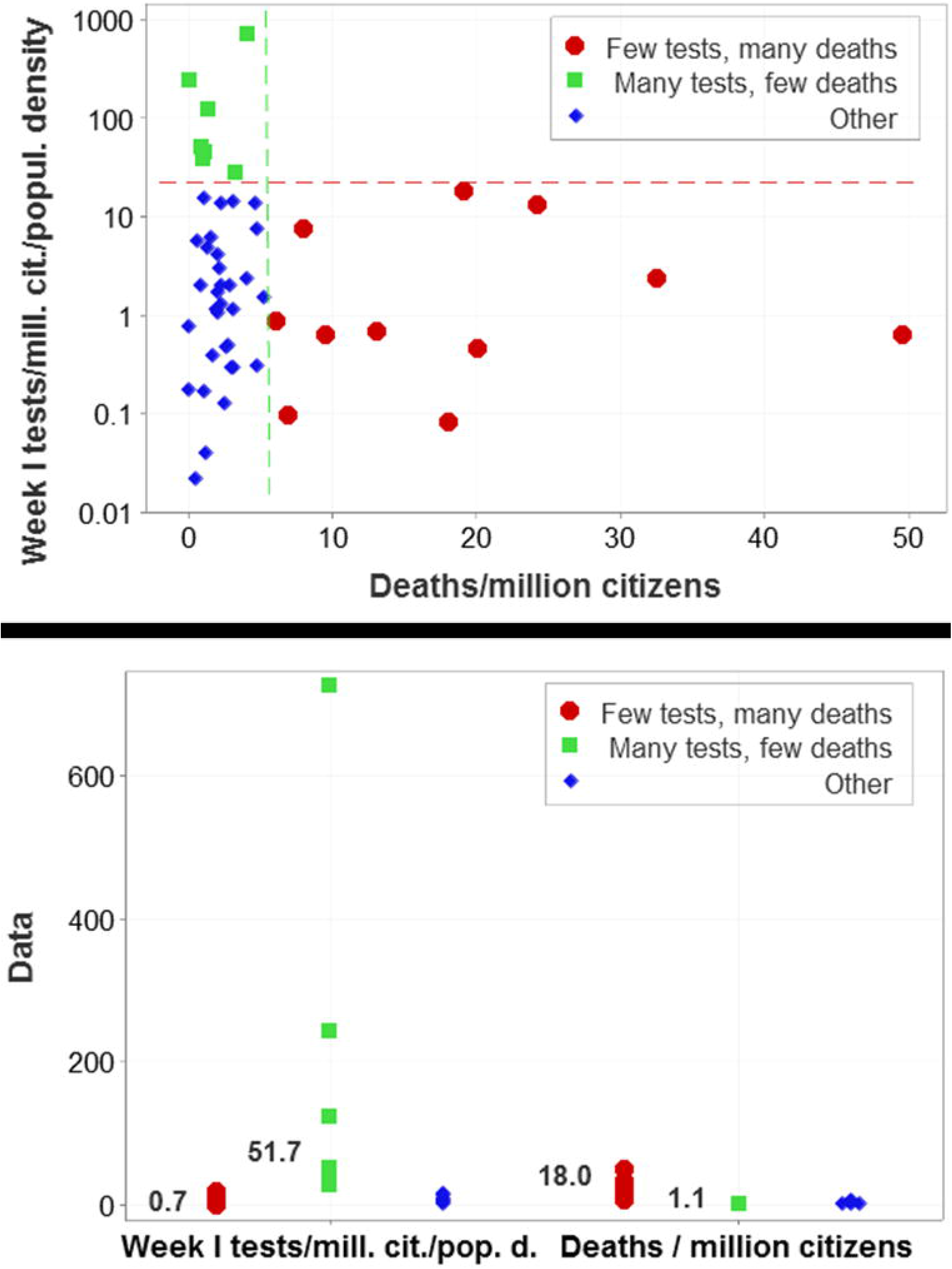
Recognition of COVID-19 mortality-related patterns across the United States. Even a small number of variables, such as the ones reported in Supplemental Table 1, provide a combinatorial template that can reveal additional relationships. For example, when the number of tests performed in the first week of testing (adjusted to state-specific population, expressed per million citizens, and also adjusted to state-specific population density) was plotted against the number of deaths per million citizens reported throughout March of 2020, two perpendicular data segments were observed and the upper limit of each data segment was used as a threshold, that is, the intersection of two perpendicular lines differentiated three groups of states (vertical and horizontal lines, **A**): (i) eleven states that conducted a low number of tests in week I and reported a high number of deaths, (ii) seven states that displayed the opposite pattern, and (iii) the remaining states (**A**). The medians of the variables analyzed differed ≥16 times between the group of states characterized by conducting many tests and reporting few deaths and the group of states that exhibited the opposite pattern (*p*<0.001, inserts, **B**).

Whether cases or fatalities are considered, findings indicate that reporting COVID-19 data as counts is not as informative as reporting metrics that consider two or more interacting quantities, such as the apparent prevalence rate and the number of deaths/million citizens. While isolated metrics –e.g., counts– ignore dynamics as well as geographical factors (including population density), composite metrics integrate numerous dimensions that facilitate geographically-specific interventions [3].

Although the KRLS regression method is a powerful and flexible approach to modeling predictive associations, to rapidly generate results, here it was used to only provide a snapshot-like assessment. If shorter time intervals were used, the KRLS approach could capture epidemic dynamics.

As evidenced by our nonparametric regression results, the variables analyzed offer a combinatorial template that highlights the importance of investigating metrics consisting of interacting quantities. For example, a recombination of those variables (the number of tests performed in week I/million citizens/population density) empirically demonstrate that massive and early testing may save lives (Figs. 1A and **B**). Such a finding is likely to also be influenced by several factors, including, but not limited to (i) availability of diagnostic kits, equipment, reagents, and trained personnel, (ii) availability of hospital beds and/or Intensive Care Units, and (iii) local and regional demographic and geographical interactions. For example, regions with a higher population density (more abundant and closer contacts among infected and susceptible citizens) tend to be associated with a higher connectivity (more highways, ports and/or airports), which foster epidemic spread [3].

While composite metrics could address pandemics as a group of local and regional interacting processes, the COVID-19 related information currently found in the press as well as national and international governmental agencies tends to lack point-based (high-resolution), geo-referenced information. While surface-based data are usually provided (e.g., state--related data), this type of data is an aggregate of geographical points and lines and, consequently, internal processes –those affecting specific cities or neighborhoods– are missed [10]. To better identify when and where interventions are most effective, point- (city- or neighborhood-related) and line-based (road-related) data are needed. To optimize these approaches and reduce the COVID-19 related mortality, the collection and reporting of high-resolution, geo-temporal data constructed as interactions is recommended.

## Data Availability

Supplementary data is available and attached.

## Acknowledgements

The authors appreciate the data gathering efforts of those citizens who contributed to Covid-19 tracking (https://covidtracking.com) and the comments provided by Dr. Ravi Durvasula (Loyola University Medical School-Chicago).

## Author contributions

JBH and ALR designed the study. ALH, RP, PK, SDS and FOF extracted the data and reviewed the literature. JBH conducted the statistical analyses. All authors contributed to writing of the report.

## Supplemental Files

**Table 1.** Epidemic data collected in all states of the US in March, 2020.

| State | *Tests*<br>*wk I* | *Total*<br>*tests* | *Wk I/*<br>*all tests* | *Pop*<br>*(mill)* | *Pop*<br>*dens* | *Tested/*<br>*mill* | *Cases* | *Cases/*<br>*mill inh* | *Deaths*<br>*(count)* | *Deaths/*<br>*mill* |
| --- | --- | --- | --- | --- | --- | --- | --- | --- | --- | --- |
| New York | 1661 | 172360 | 0.96 | 19.440 | 137.5 | 8866.2 | 59513 | 3061.37 | 965 | 49.63 |
| New Jersey | 284 | 35602 | 0.79 | 8.936 | 395.5 | 3984.1 | 13386 | 1497.99 | 161 | 18.01 |
| California | 4260 | 90657 | 4.69 | 39.937 | 94.2 | 2270.0 | 5708 | 142.93 | 123 | 3.07 |
| Washington | 4415 | 59206 | 7.45 | 7.797 | 42.2 | 7593.4 | 4310 | 552.78 | 189 | 24.24 |
| Michigan | 274 | 17379 | 1.57 | 10.045 | 40.1 | 1730.1 | 5486 | 546.14 | 132 | 13.14 |
| Florida | 1380 | 48998 | 2.81 | 21.992 | 129.1 | 2228.0 | 4950 | 225.08 | 59 | 2.68 |
| Illinois | 1622 | 27762 | 5.84 | 12.659 | 84.4 | 2193.1 | 4596 | 363.06 | 66 | 5.21 |
| Massachusetts | 171 | 39066 | 0.43 | 6.976 | 255.2 | 5600.1 | 4955 | 710.29 | 48 | 6.88 |
| Louisiana | 368 | 27871 | 1.32 | 4.645 | 34.2 | 6000.2 | 3540 | 762.11 | 151 | 32.50 |
| Georgia | 48 | 12596 | 0.38 | 3.991 | 25.9 | 3156.1 | 2683 | 672.26 | 80 | 20.04 |
| Pennsylvania | 403 | 33455 | 1.20 | 12.820 | 107.4 | 2609.6 | 3394 | 264.74 | 38 | 2.96 |
| Colorado | 938 | 14470 | 6.48 | 5.845 | 21.7 | 2475.6 | 2307 | 394.70 | 47 | 8.04 |
| Texas | 48 | 25760 | 0.18 | 29.472 | 42.4 | 874.0 | 2552 | 86.59 | 34 | 1.15 |
| Connecticut | 551 | 11900 | 4.63 | 3.563 | 248.2 | 3339.9 | 1993 | 559.36 | 34 | 9.54 |
| Tennessee | 73 | 20574 | 0.35 | 6.897 | 63.2 | 2983.0 | 1537 | 222.85 | 7 | 1.01 |
| Ohio | 148 | 20665 | 0.71 | 11.747 | 101.2 | 1759.2 | 1653 | 140.72 | 29 | 2.46 |
| Wisconsin | 259 | 17662 | 1.46 | 5.851 | 34.5 | 3018.6 | 1112 | 190.05 | 13 | 2.22 |
| N. Carolina | 17 | 19072 | 0.08 | 10.611 | 76.1 | 1797.3 | 1167 | 109.98 | 5 | 0.47 |
| Indiana | 149 | 9830 | 1.51 | 6.745 | 71.5 | 1457.4 | 1514 | 224.46 | 32 | 4.74 |
| Arizona | 373 | 13872 | 2.68 | 7.378 | 25.0 | 1880.2 | 919 | 124.56 | 17 | 2.30 |
| S. Carolina | 97 | 3789 | 2.56 | 5.210 | 62.8 | 727.3 | 774 | 148.56 | 16 | 3.07 |
| Oklahoma | 206 | 1634 | 12.60 | 3.954 | 21.8 | 413.2 | 429 | 108.50 | 16 | 4.04 |
| N Hampshire | 232 | 5396 | 4.29 | 1.371 | 56.6 | 3935.8 | 258 | 188.18 | 3 | 2.18 |
| N Mexico | 488 | 11179 | 4.36 | 2.096 | 5.9 | 5333.5 | 237 | 113.07 | 2 | 0.95 |
| Delaware | 129 | 3701 | 3.48 | 0.982 | 152.3 | 3768.8 | 236 | 240.33 | 6 | 6.11 |

**Table 1. Epidemic data collected in all states of the US in March, 2020 (cont'd)**
| State | *Tests*<br>*wk I* | *Total*<br>*tests* | *Wk I/*<br>*all tests* | *Pop*<br>*(mill)* | *Pop*<br>*dens* | *Tested/*<br>*mill* | *Cases* | *Cases/*<br>*mill inh* | *Deaths*<br>*(count)* | *Deaths*<br>*/ mill* |
| --- | --- | --- | --- | --- | --- | --- | --- | --- | --- | --- |
| Maryland | 447 | 13593 | 3.28 | 6.083 | 189.3 | 2234.6 | 1239 | 203.68 | 10 | 1.64 |
| Nevada | 262 | 10534 | 2.48 | 3.139 | 10.9 | 3355.8 | 920 | 293.08 | 15 | 4.77 |
| Missouri | 236 | 14107 | 1.67 | 6.169 | 34.2 | 2286.7 | 903 | 146.37 | 12 | 1.94 |
| Virginia | 314 | 10609 | 2.95 | 8.626 | 77.9 | 1229.9 | 890 | 103.17 | 22 | 2.55 |
| Alabama | 352 | 5014 | 7.02 | 4.908 | 36.1 | 1021.6 | 830 | 169.11 | 4 | 0.81 |
| Mississippi | 969 | 3318 | 29.20 | 2.989 | 23.8 | 1110.1 | 758 | 253.59 | 14 | 4.68 |
| Utah | 279 | 13993 | 1.99 | 3.282 | 14.9 | 4263.6 | 719 | 219.07 | 2 | 0.60 |
| Oregon | 1023 | 11426 | 8.95 | 4.301 | 16.9 | 2656.6 | 548 | 127.41 | 13 | 3.02 |
| Minnesota | 889 | 17657 | 5.03 | 5.700 | 25.3 | 3097.7 | 503 | 88.24 | 9 | 1.57 |
| Kentucky | 207 | 6018 | 3.43 | 4.499 | 43.0 | 1337.6 | 439 | 97.57 | 9 | 2.00 |
| Arkansas | 115 | 3453 | 3.33 | 3.038 | 22.1 | 1136.6 | 426 | 140.22 | 6 | 1.97 |
| Iowa | 330 | 5349 | 6.16 | 3.179 | 21.8 | 1682.6 | 336 | 105.69 | 4 | 1.25 |
| Kansas | 167 | 4513 | 3.70 | 2.910 | 13.7 | 1550.9 | 319 | 109.62 | 6 | 2.06 |
| Idaho | 427 | 4706 | 9.07 | 1.826 | 8.4 | 2577.2 | 310 | 169.77 | 6 | 3.28 |
| R. Island | 550 | 3134 | 17.54 | 1.056 | 263.9 | 2967.8 | 294 | 278.40 | 3 | 2.84 |
| Maine | 272 | 3647 | 7.45 | 1.345 | 14.7 | 2711.5 | 253 | 188.10 | 3 | 2.23 |
| Vermont | 291 | 3701 | 7.86 | 0.628 | 25.2 | 5893.3 | 235 | 374.20 | 12 | 19.10 |
| Hawaii | 12 | 8013 | 0.14 | 1.412 | 49.9 | 5674.9 | 175 | 123.93 | 0 | 0 |
| Montana | 160 | 4069 | 3.93 | 1.086 | 2.8 | 3746.8 | 161 | 148.25 | 1 | 0.92 |
| W. Virginia | 38 | 3108 | 1.22 | 1.778 | 28.3 | 1748.0 | 124 | 69.74 | 0 | 0 |
| Nebraska | 272 | 2345 | 11.59 | 1.952 | 8.9 | 1201.3 | 120 | 61.47 | 2 | 1.02 |
| Alaska | 227 | 3654 | 6.21 | 0.734 | 0.4 | 4978.2 | 114 | 155.31 | 3 | 4.08 |
| N. Dakota | 393 | 3724 | 10.55 | 0.761 | 4.2 | 4893.6 | 98 | 128.77 | 1 | 1.31 |
| S. Dakota | 185 | 3218 | 5.74 | 0.903 | 4.5 | 3563.7 | 90 | 99.66 | 1 | 1.10 |
| Wyoming | 308 | 1641 | 18.76 | 0.567 | 2.2 | 2894.2 | 87 | 153.43 | 0 | 0 |

**Table 2.** KRLS regression of potential predictors of COVID-19 related mortality.

**A – Predictors of the mortality count**
| Predictors | Estimate | Standard error | *t* value | Pr (>\|*t*\|) (*p*-value) |
| --- | --- | --- | --- | --- |
| *Tests wk I* | 0.009 | 0.010 | 0.926 | 0.36 |
| *Total tested* | 0.0002 | 0.003 | 0.650 | 0.52 |
| *Wk I / all tests (%)* | -104.35 | 118.8 | -0.878 | 0.38 |
| *Tested/mill* | -0.004 | 0.003 | -1.153 | 0.25 |
| *Cases* | 0.002 | 0.0007 | 3.430 | <0.01 |
| *Cases/mill inh* | 69104 | 21607.2 | 3.198 | <0.01 |

**B – Predictors of deaths per million citizens**
| Predictors | Estimate | Standard error | *t* value | Pr (>\|*t*\|) (*p*-value) |
| --- | --- | --- | --- | --- |
| *Tests wk I* | 0.00061 | 0.0009 | 0.611 | 0.54 |
| *Total tested* | <-0.0001 | <0.0001 | -1.314 | 0.19 |
| *Wk I / all tests (%)* | 1.99 | 11.1 | 0.179 | 0.86 |
| *Tested/mill* | 0.0002 | 0.0003 | 0.773 | 0.44 |
| *Cases* | 0.0001 | <0.0001 | 2.214 | 0.03 |
| *Cases/ mill inh* | 15623.3 | 2017.1 | 7.746 | <0.0001 |

Estimates are sample-average partial derivatives. Predictors accounted for 86.7% of the variance in deaths per million citizens ( $R^2$ : 0.8675).

